# Disrupted Bilateral Coordination of Soleus Motor Units during Early Subacute Stroke Rehabilitation

**DOI:** 10.64898/2025.12.18.25342574

**Authors:** Jackson T. Levine, Xin S. Yu, Alyssa Jones, Rebecca Muñoz, Martin Zaback, Christopher K. Thompson, Dario Farina, Simon Avrillon, José L. Pons

**Affiliations:** Northwestern University, McCormick School of Engineering, Evanston, IL 60208 USA; Shirley Ryan AbilityLab, Chicago, IL 60611 USA; Northwestern University, Department of Physical Medicine and Rehabilitation Evanston, IL 60208 USA; University of Memphis, Doctor of Physical Therapy Program, College of Health Sciences, Jackson, TN 38301 USA; University of Illinois Chicago, Department of Physical Therapy, Chicago IL, 60607 USA; Temple University, Department of Health and Rehabilitation Sciences, Philadelphia, PA 19122; Imperial College London, Department of Bioengineering, London, SW7 2AZ UK; Nantes Université, Movement-Interactions-Performance (MIP), UR 4334, Nantes, France

**Keywords:** stroke, balance, rehabilitation, motor units, synchronization

## Abstract

Stroke can affect sensorimotor control, impairing balance and locomotion. These impairments increase fall risk, limit patient independence, and reduce quality of life. Here, we investigated how stroke affects bilateral coordination of soleus motor units during standing in individuals undergoing subacute rehabilitation. Fourteen individuals were recruited after admission for inpatient rehabilitation with sixteen age- and sex-matched controls. Both groups attended two sessions separated by one week, during which high-density electromyography (EMG) signals were recorded from soleus muscles while standing on force plates. Patients also performed the Berg Balance Scale (BBS). EMG signals were decomposed into motor unit spike trains, from which peristimulus time histograms and EMG waveform averages were computed in relation to peaks in center of pressure (COP). The amplitude of motor unit and EMG activity around COP peaks, directional tuning, and bilateral synchronization were estimated. Individuals post-stroke improved their BBS score between visits, but COP displacement and speed were still greater than controls. Soleus activity was tuned anteriorly in both control and non-paretic limbs, while it was tuned laterally in paretic limbs. Synchronization of soleus activity was also reduced in the paretic limb compared to controls. Furthermore, cross-correlation lag between limbs was greater in individuals post-stroke than in controls at visit 1, while it decreased to control levels at visit 2 (*P*=0.008). The decrease in lags was strongly correlated with improvement in BBS score (*R*=-0.94, *P*<0.001). These results highlight the importance of bilateral motor unit coordination in balance, which is disrupted in both spatial and temporal domains following stroke.

**Key Points Summary:**

- We examined bilateral motor unit coordination during quiet standing in individuals in the early subacute phase following unilateral stroke, while previous works have focused on the chronic phase post-stroke.
- While the non-paretic limb of patients exhibited a spatial tuning of its activity oriented in the same direction as those of neurologically intact controls, the spatial tuning of the activity of the paretic limb was altered.
- Bilateral synchronization between soleus motor units was reduced in individuals post-stroke in comparison with neurologically intact controls.
- The level of bilateral synchronization between soleus motor units was associated with balance function and improved after one week of inpatient rehabilitation.

## Introduction

Stroke is a neurological deficit resulting from an acute focal injury to the central nervous system caused by a vascular event (Campbell & Khatri, 2020). This injury dramatically impairs cognitive and sensorimotor functions, with, for instance, half of stroke survivors exhibiting balance impairments (Khan & Chevidikunnan, 2021). These impairments are characterized by greater sway variability and greater speeds (Mizrahi *et al*., 1989; Dickstein & Abulaffio, 2000; Garland *et al*., 2003; Garland *et al*., 2007), primarily resulting from pathological neural activation of postural muscles and their coordination. Garland and colleagues (2003; 2007), for example, identified a reduced activation of the soleus muscle in the non-paretic limb using electromyographic (EMG) recordings, with fewer differences between limbs observed following rehabilitation. Furthermore, investigations into the bilateral coordination during standing balance have revealed reduced synchronization between limbs in individuals post-stroke (Arya & Pandian, 2014; Dietz & Schrafl-Altermatt, 2016; Yamanaka *et al*., 2023).

These changes underlie alterations in the activation and coordination of motor units – the neural structures that convert neural signals into motor outputs. In neurologically intact individuals, motor units that serve the same function receive a high degree of common synaptic input from both supraspinal and spinal circuits (Castronovo *et al*., 2015; Negro *et al*., 2016; Thompson *et al*., 2018), constraining them to coordinated activation and motor output. During unperturbed standing balance tasks in neurologically intact individuals, motor units from the soleus are tonically activated for weight support and show some degree of rate modulation. Motor units from gastrocnemius medialis are recruited intermittently during larger balance adjustments and gastrocnemius lateralis exhibit little activation (Vieira *et al*., 2012; Héroux *et al*., 2014; Pollock *et al*., 2014; Hug *et al*., 2021b; Cohen *et al*., 2023b). Motor unit activation varies with the magnitude and direction of postural sway, with soleus motor units demonstrating the greatest modulation with center of pressure (COP) movements along the anteroposterior axis (Cohen *et al*., 2023a; Zaback & Thompson, 2026). Bilateral motor unit synchronization is present during balance tasks in neurologically intact individuals (Gibbs *et al*., 1995; Boonstra *et al*., 2009).

In individuals post stroke, motor units in the paretic limb exhibit reduced discharge rates, compressed rate coding, and altered recruitment patterns, and the extent of these changes are moderately correlated with functional impairments (Kallenberg & Hermens, 2011; Chou *et al*., 2013; McNulty *et al*., 2014; Miller *et al*., 2014b; Mottram *et al*., 2014; Hu *et al*., 2015; Li *et al*., 2015; Hu *et al*., 2016; Negro *et al*., 2020; Beauchamp *et al*., 2023; Tacca *et al*., 2026). During standing balance, a reduced correlation between bilateral motor unit discharge rates has been observed in individuals with chronic hemiparetic stroke (Garland *et al*., 2014), suggesting they may arise from long-term adaptation in motor unit properties or numbers (Lukács, 2005; Li *et al*., 2011), or from the reorganization of synaptic inputs to these motor pools (Karbasforoushan *et al*., 2019; Beauchamp *et al*., 2023). Whether these changes occur during the acute phase of stroke, or emerge only over time, remains to be determined.

Here, we aimed to compare the activity of motor unit pools during a quiet standing balance task in patients in the early subacute phase post-stroke and in neurologically intact controls. We conducted two experimental sessions one week apart during inpatient rehabilitation. We specifically investigated soleus motor unit activation during COP peaks, focusing on the amplitude and directional tuning, as well as bilateral coordination of motor units. We hypothesized that individuals post-stroke would exhibit greater differences in directional tuning between legs, particularly in the paretic limb due to previously reported weakened neural drive (Kallenberg & Hermens, 2011; McNulty *et al*., 2014). Accordingly, we also hypothesized that the bilateral coordination of motor units would be reduced in individuals post-stroke compared to controls. Finally, we hypothesized that motor unit bilateral coordination in individuals post-stroke would be related to the level of impairments quantified with the Berg Balance Scale (BBS).

## Materials and methods

### Ethical approval and participants

14 individuals with subacute stroke admitted for inpatient rehabilitation were recruited for this study (time since stroke to first session: 19±8 days, *n*=7 females; age=60.2±15.9 years, height=171.0±6.1 cm, weight=77.6±16.3 kg, Table 1). The sample size for this study was determined based on feasibility and was consistent with prior work [14 neurologically intact participants (Zaback & Thompson, 2026)]. In the present study, we recruited a comparable number of participants post-stroke (14). The sample size is also consistent with previous motor unit studies in chronic stroke populations (Kallenberg & Hermens, 2009; Garland *et al*., 2014; Mottram *et al*., 2014; Hu *et al*., 2015; Li *et al*., 2015; Hu *et al*., 2016; McManus *et al*., 2017; Murphy *et al*., 2018; Negro *et al*., 2020).

**Table 1:** Participant Demographics.

| Stroke Participant | Age (yrs) | Sex | Stroke Type | Stroke Location | Hemi Side | Time Stroke to Admission (Days) | Time Stroke to Session 1 (Days) | Baseline BBS (out of 56) |
| --- | --- | --- | --- | --- | --- | --- | --- | --- |
| 1 | 62.0 | Male | Ischemic | MCA | Left | 16 | 24 | 48 |
| 2 | 75.7 | Female | Ischemic | MCA | Right | 6 | 18 | 28 |
| 3 | 71.1 | Male | Ischemic | Thalamus | Right | 4 | 11 | 34 |
| 4 | 56.6 | Female | Hemorrhagic | BG | Left | 10 | 15 | 29 |
| 5 | 19.0 | Male | Hemorrhagic | FP | Left | 10 | 15 | 5 |
| 6 | 75.8 | Male | Ischemic | PCA | Right | 4 | 10 | 30 |
| 7 | 67.2 | Female | Ischemic | MCA | Right | 17 | 26 | 37 |
| 8 | 65.8 | Male | Ischemic + Hemorrhagic | MCA | Right | 7 | 35 | 24 |
| 9 | 58.9 | Female | Hemorrhagic | PLIC | Right | 5 | 10 | 19 |
| 10 | 55.3 | Female | Hemorrhagic | BG | Left | 20 | 27 | 20 |
| 11 | 78.9 | Male | Ischemic | FP | Right | 5 | 12 | 34 |
| 12 | 55.4 | Male | Ischemic + Hemorrhagic | MCA | Right | 12 | 26 | 43 |
| 13 | 37.9 | Female | Ischemic | MCA | Right | 28 | 30 | 38 |
| 14 | 63.1 | Female | Ischemic | PLIC | Left | 9 | 13 | 9 |
| **Avg** | **60.2** | **M/F** |  |  | **R/L** | **10.93** | **19.00** | **28.00** |
| **± Std** | **±15.9** | **7/7** |  |  | **9/5** | **±7** | **±8** | **±12** |

*Abbreviations: Middle Cerebral Artery (MCA), Posterior Cerebral Artery (PCA), Posterior Limb of Internal Capsule (PLIC), Basal Ganglia (BG), Fronto-parietal (FP).*

16 age- and sex-matched controls (*n*=8 females, age=61.2±14.7 years) were also recruited. Two additional controls were recruited to increase the likelihood of decomposing sufficient motor units to establish normative levels of motor unit function. Individuals post-stroke were included upon meeting the following criteria: 1) first stroke (ischemic or hemorrhagic); 2) supratentorial stroke location; 3) study completion within first six weeks post-stroke; 4) currently admitted to inpatient rehabilitation and enrolled throughout the entirety of the study (at least one week from baseline assessment); 5) score of less than or equal to 1 on question 1b (i.e., asking what month it is and the age of the participant) and 0 on question 1c (i.e., asking participant to open/close eyes or grip/release the therapist’s hand) of NIH Stroke Scale for cognitive function (Brott *et al*., 1989); 6) baseline visible muscle contraction of ankle plantarflexors; 7) no history of other neurological condition or injury that would impact motor recovery; and 8) no concurrent participation in other lower limb research studies that could confound results. The institutional review board of Northwestern University approved the protocol of this study (STU00214290), which followed the standards of the declaration of Helsinki. Participants (or a power of attorney) provided their informed written consent before baseline assessment. Consent and baseline assessment were completed in the first session for all participants. The second session replicated the baseline assessment one week later. All patients received standard physical therapy throughout the study as dictated by their clinicians. This consisted of a standard of 180 minutes of skilled therapy per day for five to six days per week, distributed between physical therapy (with a focus on high-intensity gait training), occupational therapy, and speech therapy.

### Protocol

Participants stood on a split-belt treadmill (Woodway, USA) instrumented with force plates while standing shoulder-width apart with both feet aligned anteriorly and eyes open or eyes closed. Each trial lasted 30 seconds. All participants wore a harness for safety that did not provide support to the participant during standing, and a physical therapist provided assistance as necessary. The level of assistance was recorded for each task by an experienced physical therapist on the following scale: independent (0), supervision (1), touch assistance/supervision (2), touch assistance (3), partial assistance (4), and substantial assistance (5). Ground reaction forces were recorded from the instrumented treadmill at a sampling rate of 1500 Hz using a 16-bit A-D converter. Force and EMG data were synchronized using an external trigger (Micro1401, CED, UK) sent to both acquisition systems (EMG-Quattrocento; 400-channel EMG amplifier, OT Bioelettronica, Italy).

The BBS was recorded weekly as a part of standard clinical care, and results from the clinical assessments were obtained for this study.

### Electromyographic Recording

Surface EMG signals were recorded from the soleus muscle of both limbs using two-dimensional adhesive grids of 64 electrodes (GR08MM1305, 13×5 gold-coated electrodes with one electrode absent on a corner; interelectrode distance: 8mm, OT Bioelettronica, Italy). The grids were positioned lateral to the Achilles tendon in the muscle shortening direction. The skin was shaved and cleaned with an abrasive pad and an alcohol wipe prior to placing electrodes. A disposable adhesive foam layer held the grid of electrodes on the skin, and its cavities were filled with conductive paste to improve the skin-electrode contact. Reference electrodes (Ag/AgCl snap electrodes, Noraxon, USA) were placed over the patella, and a wetted strap ground electrode was placed around the ankle. The EMG acquisition system recorded signals in a monopolar montage at a sampling rate of 2048 Hz.

### Data Analysis

#### Center of Pressure

The COP displacement was computed along the anteroposterior and mediolateral axes from the split-belt treadmill and low pass filtered with a third order Butterworth filter at 5Hz. Root mean square (RMS) COP displacement and COP speed were computed for the anteroposterior and mediolateral directions as measures of balance variability and total displacement.

#### Bipolar EMG

EMG signals from each channel were visually inspected offline. Channels with significant artifacts due to movements or disrupted electrode-skin contact were manually excluded from further analyses. Across control and stroke participants, an average of 1.5 and 1.9 channels out of 64 were removed, respectively. Bipolar EMG signals were computed by taking the differential of every pair of artifact-free channels along the five columns, bandpass filtering (second order Butterworth filter, 20-500 Hz cutoff frequency), and performing signal rectification. The mean of all bipolar signals across the grid was computed and used for the analyses on EMG amplitude.

#### EMG decomposition

Bandpass filtered monopolar EMG signals were decomposed into motor unit pulse trains using a blind source separation algorithm (Negro *et al*., 2016). Decomposition was run using the peel-off option, and a silhouette value of 0.85 was used as a threshold to keep the valid motor units pulse trains. Following the automatic peak detection identifying the discharge times, all motor unit pulse trains were visually inspected and manually edited to remove false positives (artifacts) and negatives (missed peaks) when necessary (Del Vecchio *et al*., 2020; Hug *et al*., 2021a; Avrillon *et al*., 2024). Motor units were considered as duplicates when they shared at least 30% of their discharge times with a tolerance of one data point (i.e., 0.5ms) (Holobar *et al*., 2014). Motor unit activity and EMG signals were then downsampled to 1500 Hz to maintain temporal alignment with force data. Muscles with less than three identified motor units were excluded from further motor unit analyses.

#### Peristimulus waveforms and directional tuning

Previous work has demonstrated that reversals in anteroposterior COP movement are accompanied by phasic changes in soleus muscle activation (Loram *et al*., 2005; Zaback *et al*., 2023). Therefore, by trigger-averaging motor unit and EMG activation in relation to prominent COP peaks, it is possible to investigate the coordination of phasic neural drive to bilateral motor unit pools in individuals post-stroke compared to neurologically intact individuals.

Anteroposterior and mediolateral COP time-series were extracted and used to segment the motor unit discharge times and EMG time-series data. In neurologically intact individuals, the standard deviation of the high-pass filtered (second order, 0.1 Hz cut-off dual-pass Butterworth) COPs was used to set an individualized threshold for identifying prominent peaks. However, as individuals post-stroke often display large balance adjustments that may skew this threshold, standard deviation was calculated across segmented high-pass filtered COP data (*window size* = 5s, *step size* = 1.25s, total of 24 segments/30s trial), and the lower 25^th^ percentile of these measures of variability was used to set an optimal peak detection threshold that would be unaffected by the presence of aberrant fluctuations.

Windows of 3s centered over the events were used to segment motor unit discharge times and EMG activity to analyze motor unit and muscle activations relative to COP peaks. Motor unit peristimulus time histograms (PSTH) were computed relative to COP peaks in anteroposterior and mediolateral positions. The PSTH was computed over 5ms bins with all motor unit firings from a muscle pooled together and normalized by the number of peaks in each direction. Only motor units that were discharging repetitively during the pre-stimulus period were included in this analysis (>3 spikes without any interspike intervals>400ms). PSTH was baseline corrected for each direction by subtracting the mean values between -1500ms and -1000ms. Similarly, EMG signals were centered around COP peaks in the anterior, posterior, medial, and lateral directions, and also baseline-corrected for each direction by subtracting the mean values between -1500ms to -1000ms. To quantify the PSTH and EMG activation amplitude during COP peaks in the four directions, the area under the curve from -400ms to 0ms relative to the time of the COP peak was computed. This time was selected to quantify the activity that resulted in the change in direction and the resulting COP peak, as done in previous works (Zaback *et al*., 2023; Zaback & Thompson, 2026).

This analysis was then extended to investigate how each motor unit pool activity is tuned with stabilizing torques oriented throughout the entire two-dimensional transverse plane. Using procedures described by Zaback and Thompson (2026), the two-dimensional COP data were iteratively rotated in 5° increments, generating a total of 72 unique signals representing the unidimensional movement of the COP across the transverse plane. The same peak detection algorithm described above was applied to each signal, and these event time stamps were used to generate 72 baseline-corrected PSTH and waveform-averaged EMG signals representing motor unit and muscle activation time-locked to stabilizing adjustment in different directions (Fig. 1). The area under the curve of the PSTH and EMG was estimated between -400ms and 0ms relative to the peak for each direction, and directional tuning curves were generated from the area values. The circular mean (Berens, 2009) of each tuning curve, which represents the direction of COP movement generally associated with the greatest muscle activation. To enable comparison between left and right limbs without introducing side bias, directional tuning curves from the left limb were reflected across the anterior–posterior axis. This transformation aligned both limbs to a common coordinate system such that anterior = 0°, posterior = 180°, medial = 270°, and lateral = 90° regardless of limb side (Fig. 1). A medial COP peak was defined as a peak directed toward the contralateral limb, whereas a lateral COP peak was defined as a peak directed away from the contralateral limb (i.e., toward the side of the limb of interest).

**Figure 1:**
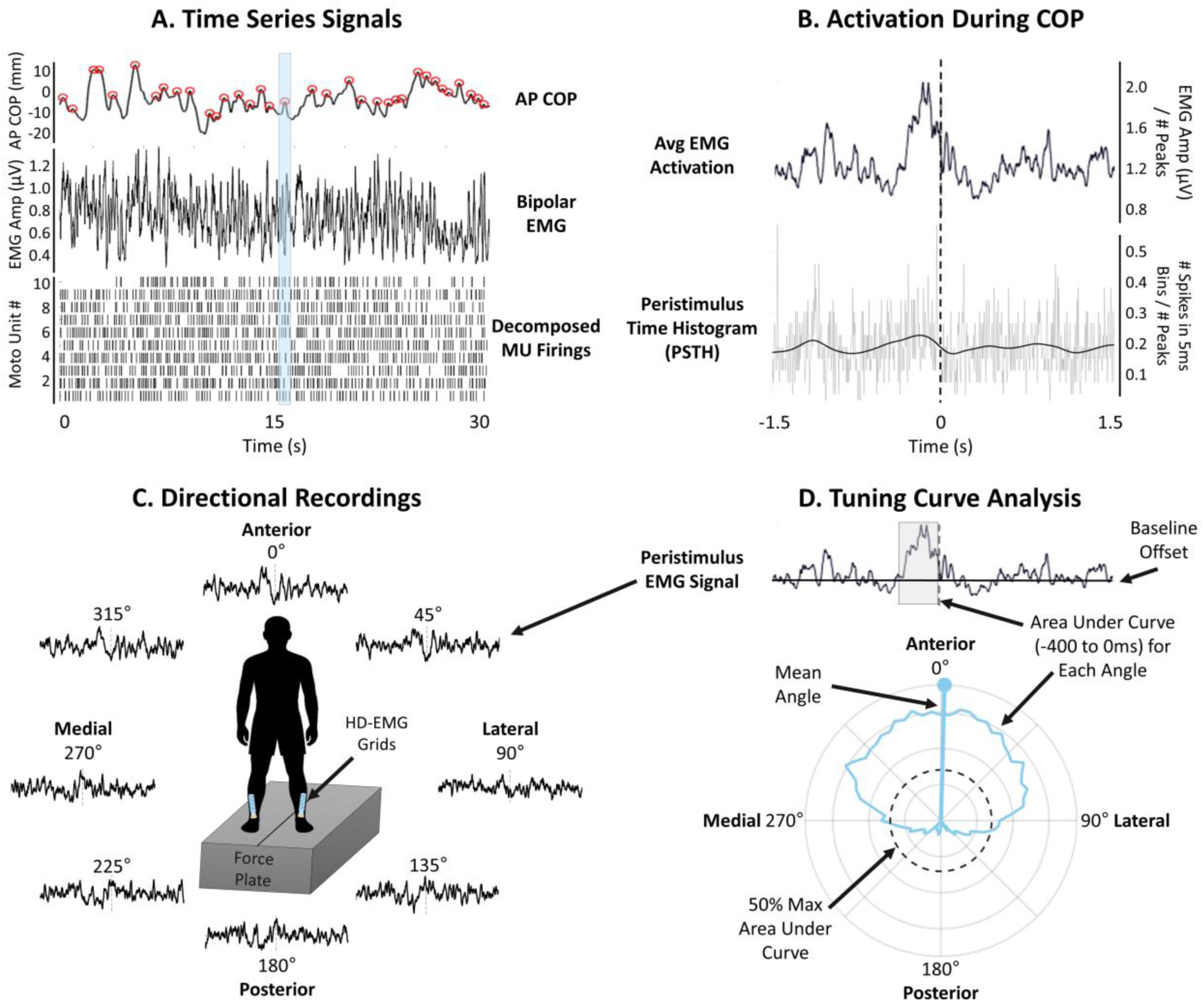
Experimental Approach. A) Anteroposterior (AP) center of pressure (COP), bipolar EMG signal, and decomposed motor unit (MU) firings were extracted during a 30s trial where participants stood on two force plates during quiet standing (as shown in C). Data were synchronized, peaks in the COP were identified, and for each COP peak, a 3s window of EMG and MU activity was extracted around the peak, as shown with the blue transparent rectangle. B) EMG and MU peristimulus time histogram (PSTH) waveform averages time-locked to COP peaks were computed. C) The time-locked waveform averages (EMG signals shown in the figure) were computed during COP peaks in the anterior, posterior, medial, and lateral directions within the transverse plane of axial rotation, as well as in 5° increments around the participant. D) Directional tuning curves were constructed across these 72 directions separated by 5° and the circular mean computed to determine the directional tuning of each muscle activity. The dashed line represents the 50^th^ percentile of the area under the curve.

#### Contralateral-triggered motor unit and EMG activation

To assess bilateral coordinated activation between soleus muscles, phasic bursts of activity were identified from the cumulative spike train and EMG time series separately for each muscle. The timing and amplitude of activity observed in each muscle were then examined relative to the phasic bursts of the contralateral muscle.

The cumulative spike train of each soleus was computed by convolving the summed spike trains of all motor units within each leg with a 400ms Hanning window, and peaks in activity were identified using the same peak detection algorithm previously described to identify prominent peaks in COP time-series data. The 40^th^ percentile of the standard deviation was used as a threshold instead of the 25^th^, utilizing a more selective criteria for the inherently greater variability of motor unit discharge and noisier EMG signals that has been reported in individuals post-stroke (Thomas *et al*., 2002). PSTH of each limb were constructed in a 3s window relative to the peaks in the smoothed cumulative spike train of each limb and baseline corrected. Similarly, rectified bipolar EMG signals were low-pass filtered (second order, 2 Hz cutoff frequency, Butterworth), and the same approach was used to identify EMG time-time series peaks and generate baseline corrected waveform averages in a 3s window relative to peaks in the smoothed EMG. As previously done, PSTH and EMG amplitude was estimated as the area under the curve in a window of -200ms to +200ms relative to the event. The difference between contralateral and ipsilateral activation amplitude (*Amplitude_contra_ – Amplitude_ipsi_*) was then computed, with a value of zero representing amplitude of contralateral-triggered activation equal to ipsilateral-triggered activation. A value less than zero represents a reduced amplitude of contralateral-triggered activation compared to ipsilateral triggered activation.

#### Bilateral cross correlation

To further assess bilateral coordination of motor unit and EMG activations, cross correlations were computed on PSTH and EMG waveform averages time-locked to anterior COP peaks (0°). All signals were baseline corrected and smoothed with a Gaussian kernel of 50ms. The maximum absolute correlation coefficient and the absolute lag at the time of the maximum absolute correlation coefficient were computed for each trial with no maximum lag enforced. To determine if there was significant coordination between limbs, the cross-correlation coefficient was compared to a bootstrapped significance threshold that was computed by shuffling the discharge times and EMG time-series and calculating the cross-correlation coefficient. The 95^th^ percentile of the bootstrapped cross correlation coefficients over 100 iterations was set as the significance threshold.

### Statistics

For all analyses but the directional-tuning analysis, we used linear mixed effects models in SPSS 26.0.0.1. Normality of the data was assessed using the Shapiro-Wilk test, as well as by visually examining skewness, kurtosis, and histograms. For non-normal distributions, square root transformations were performed (West *et al*., 1995; Kim, 2013). If the transformed data remained non-normal, a generalized linear mixed effects model was used. We compared the BBS of individuals post-stroke only across visits using a linear mixed effects model with Visit (1, 2) as a fixed effect and Participant as a random effect. We compared the RMS COP displacement (independently for different directions), COP speed (independently for different directions), and cross correlation coefficient and lag using a linear mixed effects model with Group (control, patient), Visit and Task (eyes open, eyes closed) as fixed effects and Participant and Assist level (scored 0-5) as random effect to account for the degree of independence in the tasks. We compared the amplitude of PSTH and EMG independently for each direction, as was done with the COP metrics, with Limb (control right, control left, paretic, non-paretic), Visit, and Task as fixed effects and Participant and Assist level as random effects. The random structure of all models included random slopes and intercepts to also account for between participant variability in the fixed effects. Ultimately, the model associated with the lowest Bayesian information criterion was retained for the analysis.

Circular statistical analyses were performed on the circular mean angle of PSTH and EMG using the circular statistics MATLAB toolbox (Berens, 2009). The Harrison-Kanji test, i.e., the equivalent of a two-way ANOVA for circular data, was used to identify significant effects of Limb, Visit, and Task. All combinations of two-way interaction were evaluated separately, as no three-way circular ANOVA was available. Where significant interactions were identified, the Watson Williams test, i.e., the equivalent of a one-way ANOVA for circular data, was used to investigate differences within each factor with significance level of 0.05 with a Bonferroni adjustment (i.e., α/*k*, where k is the number of contrasts).

We compared the difference in amplitude of contralateral- and ipsilateral-triggered activation with a linear mixed effects model with Limb (control left/right, control right/left, non-paretic/paretic, paretic/non-paretic), Visit and Task as fixed effects and Participant and Assist level as random effects. The selection of the random structure of the models followed the same procedure as above. Significance level was set at 0.05. Bonferroni corrections were used in post hoc tests for pairwise comparisons and Cohen’s d effect sizes were reported with 95% confidence intervals. Outliers exceeding three standard deviations from the mean were excluded.

To determine the relationship between neuromuscular outcomes and functional balance in a stroke population, a linear regression and a repeated measures correlation were performed to examine how the change in BBS was related to the change in any outcome measure that demonstrated a significant change across visits. For this analysis, only individuals with valid data at both visits were included.

## Results

### Balance and center of pressure

Individuals post-stroke improved their BBS score from visit 1 (28.43±12.19) to visit 2 (38.57±8.71, *P<*0.001, *d=*0.957, 95% CI: [0.175, 1.739]; Fig. 2A). Twelve out of fourteen participants improved greater than the minimal clinically important difference of five points (Tamura *et al*., 2022).

**Figure 2:**
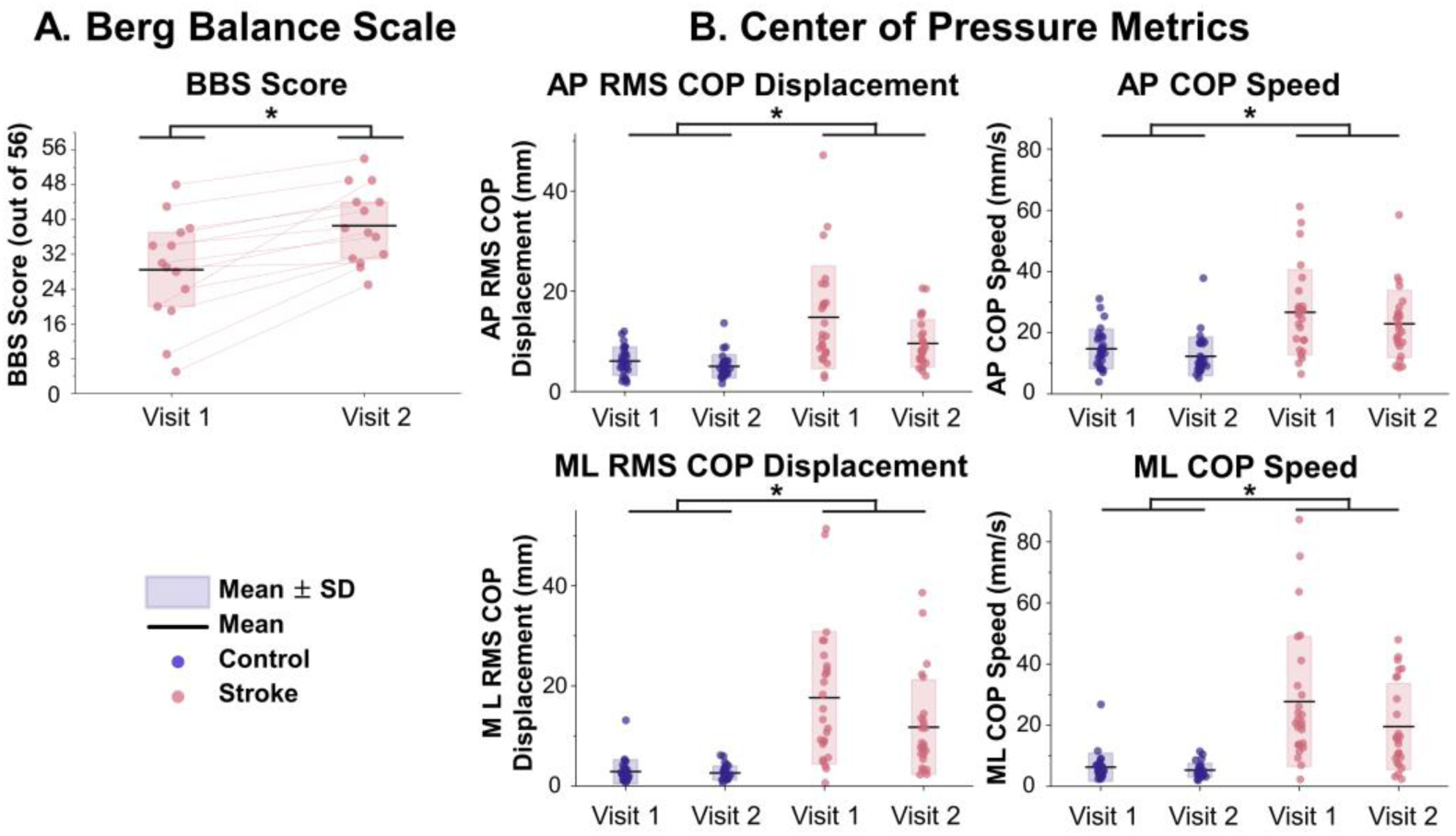
A) Berg Balance Scale (BBS) score out of 56. B) Anteroposterior (AP) and Mediolateral (ML) Root Mean Square (RMS) COP and COP Speed are displayed. * *P*<0.05.

RMS COP displacement in the anteroposterior direction was greater in the stroke group (13.48±6.44mm) compared to controls (5.53±2.60mm, *P<*0.001, *d*=0.960, 95% CI: [0.565, 1.355]) and lower at visit 2 (7.14±4.27mm) compared to visit 1 (10.27±8.53mm, *P=*0.003, *d*=0.414, 95% CI: [0.004, 0.793]). Similarly, average COP speed in this direction was greater in the stroke group (24.81±12.57mm/s) than controls (13.48±6.44mm/s, *P<*0.001, *d*=0.771, 95% CI: [0.383, 1.159]) and was lower at visit 2 (17.23±10.21mm/s) compared to visit 1 (20.50±12.21mm/s, *P=*0.009, *d*= 0.304, 95% CI: [-0.072, 0.680]).

RMS COP displacement in the mediolateral direction was greater in the stroke group (14.68±11.77mm) than controls (2.74±1.92mm, *P<*0.001, *d*=1.365, 95% CI: [0.949, 1.780]), and greater with eyes open (9.37±10.79mm) than eyes closed (7.40±9.43mm, *P=*0.009, *d*=1.693, 95% CI: [1.257, 2.128]). Average COP speed in the mediolateral direction was greater in the stroke group (23.64±18.33mm/s) than controls (5.77±3.57mm/s, *P<*0.001, *d*=1.242, 95% CI: [0.833, 1.650]), with greater speeds occurring at visit 1 (V1=16.61±18.46mm/s, V2=11.90±12.01mm/s, *P=*0.004, *d*=0.334, 95% CI: [-0.043, 0.710]). Overall, individuals post-stroke had greater COP displacement and speed than controls (Fig. 2B).

### High-density EMG decomposition

Motor unit yields are shown in Table 2. We excluded one control participant and three stroke participants from the motor unit analyses because no motor units were identified for any muscle.

**Table 2:** Motor unit yield across limbs, groups and visits.

|  |  | Stroke |  | Control |  |
| --- | --- | --- | --- | --- | --- |
|  |  | Paretic | Non-Paretic | Right | Left |
| Eyes Closed | Visit 1 | 8.8±10.2 | 5.0±6.4 | 11.1±10.7 | 9.1±9.1 |
|  | Visit 2 | 6.1±6.9 | 8.7±8.0 | 11.9±8.8 | 12.8±8.4 |
| Eyes Open | Visit 1 | 9.2±8.2 | 5.1±5.6 | 11.0±11.2 | 8.3±8.1 |
|  | Visit 2 | 6.4±7.0 | 7.0±7.2 | 13.6±9.2 | 12.3±7.4 |

### Activation amplitude during COP peaks

The amplitude of PSTH and EMG during COP peaks was computed in the anterior, posterior, medial, and lateral directions (Fig. 3). For PSTH in the lateral direction, there was an effect of limb (*P*=0.016), with the amplitude in the paretic limb (0.063±0.091 pulses/peak) greater than in the left limb of controls (0.012±0.055 pulses/peak, *P=*0.016, *d*=0.786, 95% CI: [0.309, 1.263]). No other differences were observed between limbs (all *P>*0.287). There were neither effects nor interactions in other directions (*P>*0.111).

**Figure 3:**
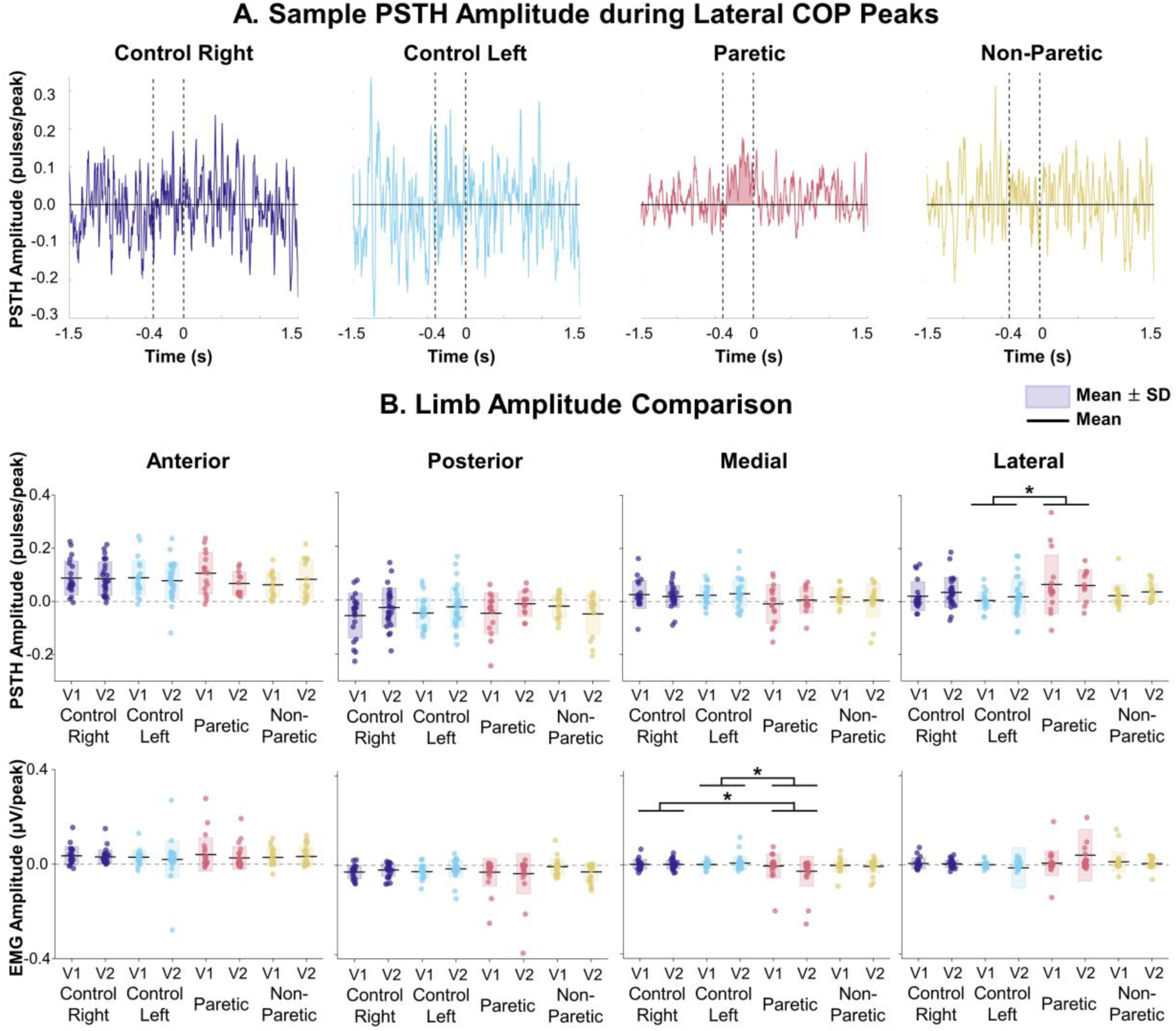
Activation amplitudes in anteroposterior and mediolateral directions. A) Sample peristimulus time histogram (PSTH) amplitude from one control and one stroke participant during lateral center of pressure (COP) peaks in the eyes closed task during visit 2. Area under the curve between -0.4s to 0s relative to the peak in COP. B) Comparison of PSTH and waveform-averaged EMG during COP peaks in anterior, posterior, medial and lateral directions. * *P*<0.05. V = Visit.

For EMG in the medial direction, there was an effect of limb (*P*=0.018), with the amplitude in the paretic limb (-0.018±0.061 μV/peak) lower than in the right limb of controls (0.002±0.020 μV/peak, *P=*0.042, *d*=0.557, 95% CI: [0.160, 0.953]) and left limb of controls (0.005±0.024 μV/peak, *P=*0.026, *d*=0.590, 95% CI: [0.185, 0.996]), but not the non-paretic limb (-0.005±0.029 μV/peak, *P=*0.113, *d*=0.448, 95% CI: [0.037, 0.860]. There were neither effects nor interactions in other direction (*P>*0.387). Collectively, these results indicate that the activity of the SOL in the paretic limb is modulated with COP displacements along the mediolateral axis.

### Directional tuning

To examine the directional tuning of the activity of the SOL, tuning curves were constructed from motor unit discharge activity (PSTH) and EMG amplitudes time-locked to COP peaks spanning the transverse plane (Fig. 4A). There was an effect of limb on the circular mean angle of these tuning curves derived from both PSTHs (*P=*0.006) and waveform-averaged EMG amplitudes (*P=*0.029). There were no effects of visit, task, nor interactions (all *P>*0.201). A Bonferroni correction was used to set the significance threshold at 0.008 (i.e., 0.05/6) for post-hoc comparisons with Watson-Williams tests.

**Figure 4:**
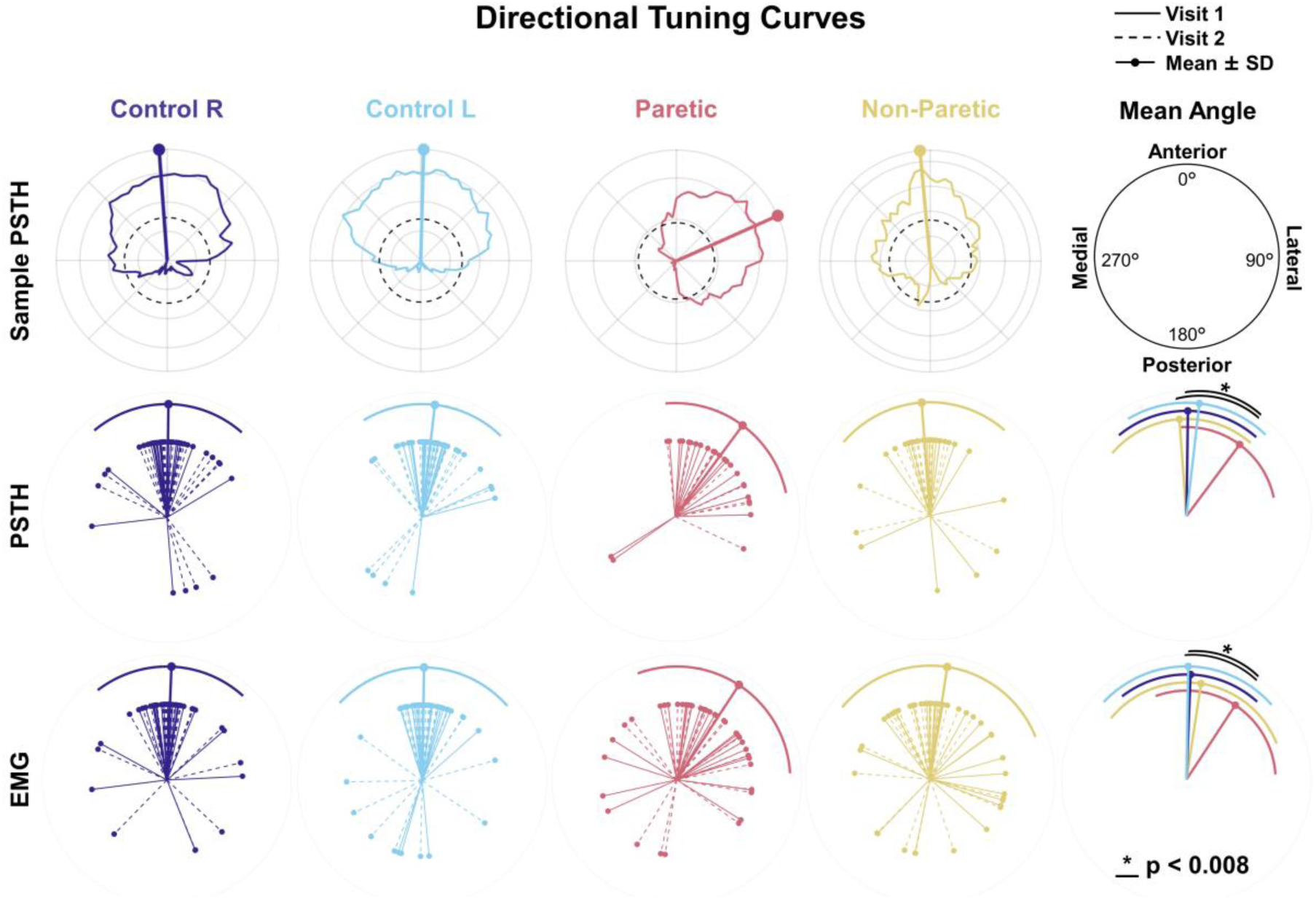
Directional tuning of soleus activity. The first row displays directional tuning curves of motor unit activity extracted from peristimulus time histogram (PSTH) from a control and a participant post stroke. Additionally, the mean circular angle of the tuning curves extracted from PSTH and EMG plots are displayed for each participant for each visit pooled across tasks, and the mean per limb is displayed. To compare rotational responses across limbs, rotations were reflected such that 90° always represents lateral tuning and 270° always represents medial tuning. * *P*<0.008.

In neurologically intact individuals, the activity of SOL in the control limbs was tuned anteriorly using both PSTH and EMG amplitude, in accordance with the previous work of (Zaback & Thompson, 2026). The same result was observed in the non-paretic limbs of individuals post-stroke. The activity of the SOL in the paretic limb, however, was tuned laterally and anteriorly (Fig. 4A). More specifically, for PSTH, the mean angle of the tuning curve was more lateral for the paretic limb (36.56±41.48°) than for the non-paretic limb (-4.17±45.39°, *P=*0.003, *d*=0.934, 95% CI: [0.400, 1.468]) and the right control limb (0.79±40.62°, *P=*0.004, *d*=0.874, 95% CI: [0.408, 1.340]). There was no difference with the left control limb (6.54±37.56°, *P=*0.012, *d*=0.770, 95% CI: [0.294, 1.247]). For EMG amplitudes, the mean angle of the tuning curve was significantly more lateral for the paretic limb (33.24±53.41°) than for the right control limb (2.27±39.90°, *P=*0.005, *d*=0.662, 95% CI: [0.267, 1.056]) and the left control limb (0.93±47.75°, *P=*0.008, *d*=0.640, 95% CI: [0.235, 1.044]). There was no difference with the non-paretic limb (8.67±59.53°, *P=*0.093, *d*=0.434, 95% CI: [0.025, 0.844]). Collectively, these results indicate that the activity of the SOL in the paretic limb was tuned laterally and anteriorly while the activity of non-paretic and control limbs were exclusively tuned anteriorly. This suggests that the greatest activation of the paretic limb occurred during lateral shifts toward the paretic limb.

### Contralateral-triggered activation

To evaluate the synchronization between limbs, we averaged PSTH and EMG amplitudes for each soleus around phasic bursts from the contralateral soleus (Fig. 5A, 5B). We then calculated the difference between contralateral- and ipsilateral-triggered activation amplitudes: zero indicates equal activation, and negative values indicate reduced activation of the limb when peaks of activity are observed in the contralateral limb (Fig. 5C).

**Figure 5:**
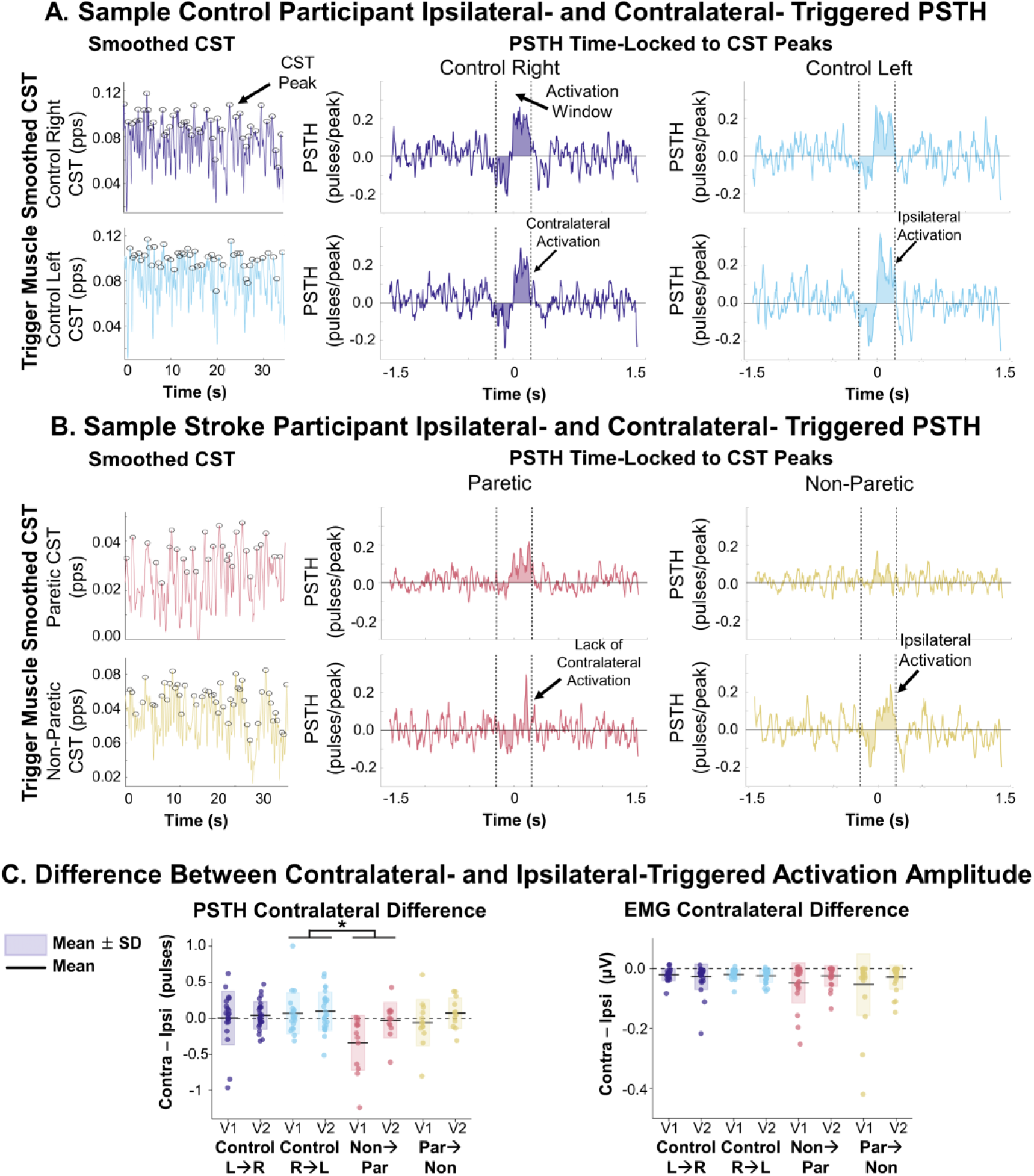
Contralateral-Triggered Activation. A) Control right (purple) and left (blue) and B) stroke participant paretic (red) and non-paretic (yellow) ipsilateral- and contralateral-triggered peristimulus time histogram (PSTH) waveforms during standing with eyes closed (visit 2 for the control and visit 1 for the stroke participant). The smoothed cumulative spike train (CST) is displayed with the peaks shown with circles. As a guide to read these figures, for the top row of Panel A, the left column is the CST of the control right limb with peaks identified. The middle column is the ipsilateral activation or activation amplitude of the right limb during right limb peaks in CST. The left column is the contralateral activation or activation amplitude of the left limb during right limb peaks in CST. C) Difference between the amplitude of the contralateral (contra) and ipsilateral (ipsi) triggered activation amplitudes. * *P*<0.05. R = Right. L = Left. Par = Paretic. Non = Non-Paretic. V = Visit. Pps = Pulses Per Second.

For PSTH, activations to contralateral peaks were different between visits (*P=*0.011) and limbs (*P=*0.004), without interactions between visits and limbs (*P>*0.199). Specifically, the difference in paretic limb activation during non-paretic limb peaks (-0.19±0.36 pulses) was lower than the difference observed in the left control limb during right limb peaks (0.08±0.27 pulses, *P=*0.002, *d*=0.734, 95% CI: [0.234, 1.234]). The differences were not significant with the non-paretic activation during paretic peaks (0.00±0.28 pulses, *P=*0.082, *d*=0.375, 95% CI: [-0.184, 0.934]), and with right control limb activation during left limb peaks (0.03±0.28 pulses, *P=*0.061, *d*=0.585, 95% CI: [0.091, 1.080]). Thus, activation amplitude of the paretic limb during phasic bursts of the non-paretic limb was substantially reduced, indicating that the paretic limb failed to coordinate reliably with the non-paretic limb. For EMG, there was no effect of visit, task, limb, nor interaction on the difference in amplitudes (*P>*0.245).

### Cross correlation of motor unit and EMG waveform averages

Cross correlation coefficients between SOL activities of each limb and the corresponding absolute lags were computed from PSTH (Fig. 6A) and EMG signals. All coefficients were deemed significant compared to the 95^th^ percentile of bootstrapped randomly shuffled signals.

**Figure 6:**
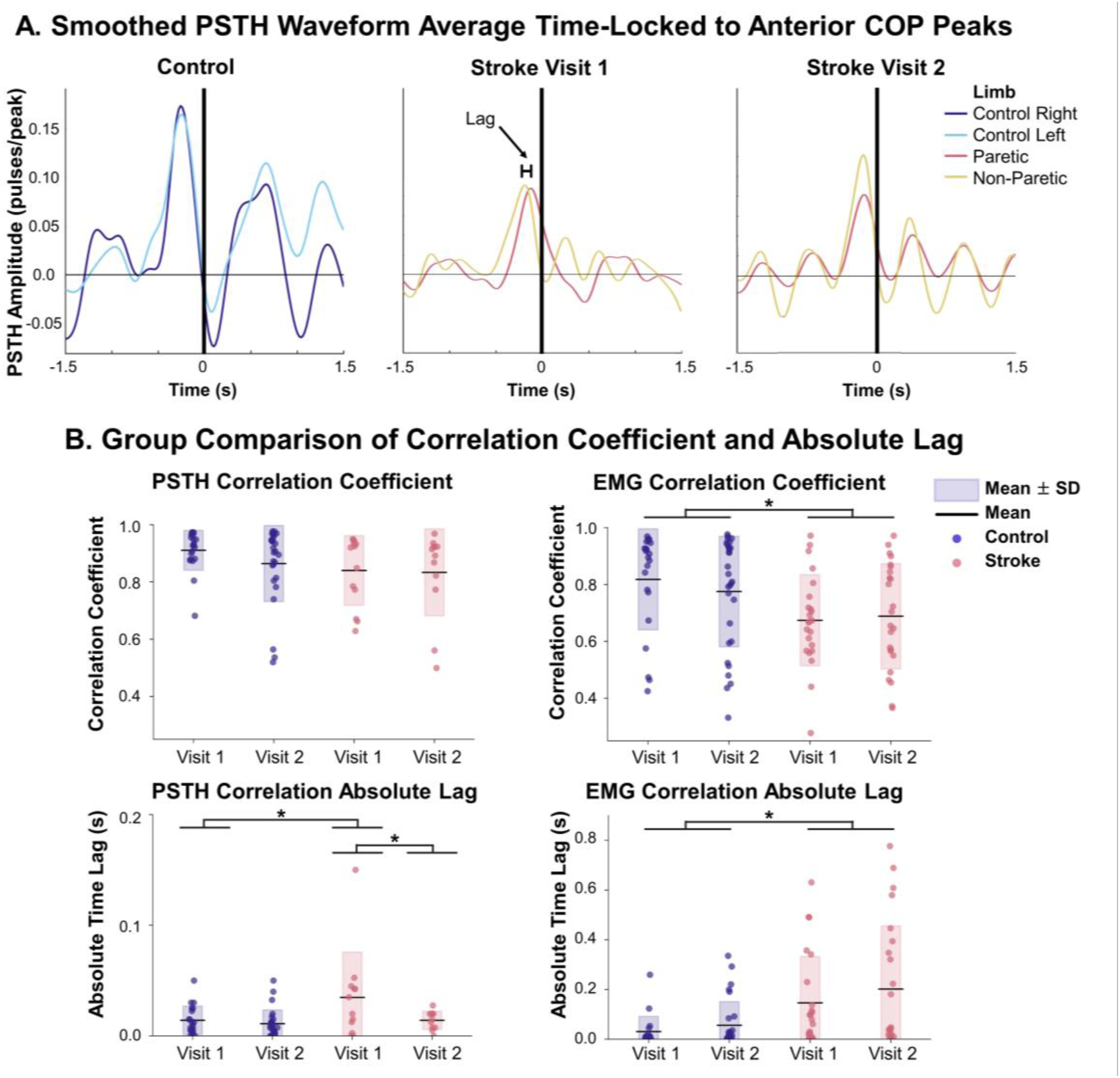
Lag between bilateral soleus activities. A) Bilateral smoothed peristimulus time histogram (PSTH) time-locked to anterior center of pressure (COP) peaks for a sample control and stroke participant from visit 1 and visit 2 during the eyes closed task. The graphical display of time lag is depicted, and the black line indicates the time of COP peak. B) Comparison of the maximum correlation coefficient and absolute time lag of smoothed PSTH and EMG between limbs. * *P*<0.05.

For PSTH, there were no effects of group, visit, task, nor interactions on maximum absolute coefficient (*P>*0.763). For absolute lag, a group*visit interaction (*P=*0.037, Fig. 6B) revealed that absolute lag was greater for the stroke group at visit 1 (0.03±0.04s) compared to visit 2 (0.01±0.01s, *P=*0.009, *d*=0.912, 95% CI: [-0.009, 1.833]) and was greater for the stroke group at visit 1 compared to controls (0.01±0.01s, *P=*0.008, *d*=4.454, 95% CI: [3.054, 5.853]). However, absolute lag of the stroke group was no longer different from controls at visit 2 (0.01±0.01s, *P=*0.491, *d*=0.319, 95% CI: [-0.438, 1.077]), while absolute lag did not differ across visits for controls (*P=*0.912, *d*=0.021, 95% CI: [-0.608, 0.651]).

For EMG, there was an effect of group (*P=*0.005, Fig. 6B), but no effects of visit, task, nor interactions on maximum absolute coefficient (*P>*0.170). Controls demonstrated greater correlation coefficients (0.79±0.19) compared to the stroke group (0.68±0.17, *P*=0.005, *d*=0.381, 95% CI: [-0.017, 0.779]). The stroke group demonstrated higher absolute lag (0.17±0.22s) than the control group (0.05±0.08s, *P=*0.001, *d*=0.916, 95% CI: [0.461, 1.371]; Fig. 6B).

Collectively, these results indicate greater lag, and therefore reduced coordination, between limbs after a stroke. Importantly, when assessing lags on activity in PSTH, the stroke group exhibited an improvement such that the bilateral synchronization improved from visit 1 to visit 2 to comparable levels to controls.

### Regressions with changes in clinical outcomes

Only PSTH absolute lag demonstrated a significant longitudinal change for stroke participants. Therefore, the relationship between the change in PSTH absolute lag and the change in functional balance score (i.e., BBS score) for stroke participants was assessed via linear regressions. Data was averaged across tasks per participant as task effect was absent. A linear regression between the changes in lag between visits and the change in BBS score between visits revealed a significant negative correlation (*R*=-0.94, *P*<0.001). This was further confirmed by a repeated measures correlation analysis with a significant negative correlation between both variables (*R*=-0.78; *P*=0.002). Thus, an increase in BBS score was accompanied by a decrease in lag (or an increase in synchronization).

## Discussion

In this study, we quantified the bilateral coordination of soleus motor units during standing balance tasks in individuals in the early subacute phase post-stroke. While clinical assessments of balance and COP displacements and speed improved between visits, individuals post-stroke continued to exhibit greater COP variability than controls. Furthermore, while control activated the soleus in coordination, both spatially along the anteroposterior axis and temporally, individuals post-stroke demonstrated differences in directional tuning and synchronization of their soleus activity. While the non-paretic limb was also activated along the anteroposterior axis, the paretic limb exhibited preferential activation along the mediolateral axis, and the two limbs in turn also lacked temporal coordination. The synchronization improved with training, and this improvement was shown to be related to improvement in functional balance, emphasizing its importance to effective balance recovery.

### Altered directional tuning of activity of the paretic limb

The soleus plays an essential role in balance, as its tonic activity with phasic modulation during weight-bearing is critical for maintaining upright stance by counteracting the gravitational torque acting on the center of mass (Loram *et al*., 2004; Di Giulio *et al*., 2009). Previous work that has examined the directional tuning of the soleus, either through analysis of activation amplitude in response to multi-directional perturbations or natural COP oscillations during unperturbed standing, has shown that the soleus activity is primarily modulated with movements of the COP or center of mass along the sagittal plane (Winter *et al*., 1993; Winter *et al*., 2003; Loram *et al*., 2005; Ting, 2007; Zaback & Thompson, 2026). Cohen and colleagues (2021; 2023a) further demonstrated that in neurologically intact individuals, some motor units in the soleus and gastrocnemius medialis respond broadly across directions, while others show direction-specific tuning. The results of this study corroborated previous findings, with motor unit activity and EMG of neurologically intact individuals directionally tuned to anterior COP peaks. During dual-legged standing, anteroposterior sway is controlled through modulation of ankle plantarflexor and dorsiflexor torques, while mediolateral sway in neurologically intact individuals is generally controlled by loading and unloading of the hip rather than by inverting and everting the ankle joints (Day *et al*., 1993; Winter *et al*., 1993; Winter *et al*., 1996). This explains the anteriorly oriented tuning of the soleus in controls. It is noteworthy that the mean angles of the tuning curve observed here exhibited higher variability than those measured by (Zaback & Thompson, 2026) (SD: 37-47° vs. 17.7°). This difference may result from differences in demographics for the control group (8 males/females, mean age: 61.2±14.7 years here vs. 10 males; mean age: 29.9±7.2 years in Zaback and Thompson, 2026), as older individuals often experience impairments in balance control (Granacher *et al*., 2011; Matsuda *et al*., 2025). Another factor explaining this variability could be the duration of trials used to construct the tuning curves, which was limited here – compared to our previous study in neurologically intact individuals – due to the inability of our patients to maintain postures for long periods of time. However, the mean angles of control and non-paretic limbs align with the previous study (Zaback & Thompson, 2026), suggesting the robustness of the method with shorter trials.

While the non-paretic limb exhibited preserved directional control with anteriorly tuned activation, the paretic limb displayed an abnormal directional tuning, exhibiting modulation of activation along the mediolateral axis (Fig. 3) and greatest activation during anterior and lateral COP movement (∼40° from anterior, Fig. 4). The abnormal modulation of paretic limb soleus activity along the mediolateral axis may be due to a couple of mechanisms. First, previous work has demonstrated slower generation of torque necessary to recover from perturbations when directed toward the paretic limb compared to perturbations directed toward the non-paretic limb or anteriorly (Palmer *et al*., 2025), indicating a weakened response on the paretic limb. The nervous system, therefore, may prioritize activation of the paretic limb only for the most destabilizing perturbations, such as lateral shifts, for which the non-paretic limb cannot fully compensate – possibly as a result of compressed rate coding, impaired recruitment, or motor unit fatigue (Kallenberg & Hermens, 2011; Chou *et al*., 2013; McNulty *et al*., 2014; Miller *et al*., 2014b; Mottram *et al*., 2014; Hu *et al*., 2015; Li *et al*., 2015; Hu *et al*., 2016; Negro *et al*., 2020; Beauchamp *et al*., 2023; Tacca *et al*., 2026). Second, individuals post-stroke exhibit impaired proprioception, sensory processing, and spinal reflexes critical to balance control and rehabilitation after stroke (Wutzke *et al*., 2013; Aman *et al*., 2014; Chia *et al*., 2019; Gorst *et al*., 2019; Johnson *et al*., 2025), which may result in abnormal directional tuning of the paretic limb. While individuals with chronic stroke exhibit increased stiffness of the gastrocnemius and soleus that could modify directional tuning and afferent feedback (Zhao *et al*., 2009; Zhao *et al*., 2015; Dias *et al*., 2019; Liang *et al*., 2024), these tissue mechanical changes may not have had enough time to develop in the acute sample recruited for this study (Lundström *et al*., 2010; Katoozian *et al*., 2018).

### Reduced bilateral synchronization post-stroke

In addition to impaired spatial coordination of motor units and EMG, disrupted bilateral temporal coordination may also help to explain the asymmetrical control of the soleus and postural instability post-stroke. Previous work has shown that anterior sway induces greater common drive between bilateral soleus muscles compared to backward sway in neurologically intact individuals (Mochizuki *et al*., 2007), while greater instability is associated with increased lag and reduced correlation between bilateral soleus muscles (Promsri, 2022). In this study, cross correlation analysis revealed greater lag between limbs during anterior COP peaks in individuals post-stroke compared to controls. Interestingly, PSTH absolute lag decreased from visit 1 to visit 2, marking the only observed longitudinal improvement in neural metrics. No other longitudinal changes were observed, which may be the result of the small sample size and the large variability between participants. The physical therapist assistance may have also impacted the outcomes, but this was accounted for in the statistical tests, and the level of assistance did not change over time (average change: -0.43±0.63 points out of 5, *P*=0.251). This change in PSTH absolute lag suggests a link between improvements in bilateral synchronization and balance recovery during early rehabilitation, but future studies with larger sample sizes will be necessary to corroborate these initial findings. Additionally, when evaluating activation amplitude during phasic peaks in the contralateral limb, there was a relative reduction in amplitude for the paretic limb compared to controls, further indicating impaired bilateral coordination.

These findings suggest potential mechanisms for impaired bilateral coordination. First, processing and transmission of proprioceptive information between limbs may be disrupted, resulting in the increased correlation lag between limbs and reduced amplitude during contralateral phasic activation. In neurologically intact individuals, when a vibratory stimulus is applied to one leg, the level of common drive between bilateral soleus muscles is reduced, suggesting that proprioceptive feedback not only plays a role in unilateral soleus modulation but also bilateral coordination (Mochizuki *et al*., 2007). Similarly, in the same study when triggering based on contralateral voluntary activation, a reduction in contralateral amplitude was observed. When triggering using contralateral non-noxious electrical nerve stimulation for individuals post-stroke, a reduction in contralateral amplitude was observed when triggering on the paretic leg due to disrupted sensory processing post-stroke (Dietz & Schrafl-Altermatt, 2016). This further supports the hypothesis that following stroke, processing of sensory information used for bilateral motor coordination is impaired, potentially impairing the ability of individuals to generate synchronous stabilizing torques.

Bulbospinal pathways, such as vestibulospinal and reticulospinal tracts, are important for modulation of soleus motor unit activity in neurologically intact individuals (Monsour *et al*., 2012), making their function critical for balance. However, a stroke may alter the bilateral gains and descending commands of these pathways, which can have debilitating effects on motor control and coordination (Marsden, 2005; Miller *et al*., 2014a; Mccall *et al*., 2017). Previous research showed that activation of brainstem pathways following stroke via galvanic vestibular stimulation led to hyperactive force responses on the non-paretic limb (Marsden, 2005), suggesting asymmetrical brainstem activation. Interestingly, it has been demonstrated that modification of balance may be mediated by a combination of central and peripheral systems, where central structures may modulate feedback loops based on task demands (Welch & Ting, 2014). Therefore, impairments in descending commands and proprioception after stroke may have contributed to the abnormal balance observed in this study. Future work is necessary to provide a more complete understanding of the neural circuits underlying impaired balance post-stroke and how they interact at the different levels.

### Motor unit coordination predictor of balance function

In addition to characterizing deficits in neural coordination during the early subacute phase post-stroke, we aimed to evaluate the relationship between changes in these neural measures and functional recovery, as assessed by licensed physical therapists using the widely used balance assessment, the Berg Balance Scale. A regression between the change in BBS and the change in synchronization of motor unit activity (PSTH absolute lag) between visits was assessed, as this was the only metric to demonstrate an improvement across visits. This analysis revealed that a change in synchronization was strongly associated with a change in BBS score (*R=-0.94*; *P*<0.001), suggesting that individuals who improve their bilateral synchronization also achieve improved functional balance.

Thus, PSTH-derived metrics may be considered as a biomarker when assessing patients’ control of balance. However, the application of such approaches is currently limited to high quality recordings capable of extracting multiple motor unit firings in patients. For example, it is noteworthy that our regression analysis only included individuals with motor units extracted in both visits, with three individuals in the stroke group excluded from all motor unit analyses due to the inability to extract any motor units. The absence of extracted motor units may be explained by physiological factors, such as low muscle activation or reduced neural drive, but also poor signal quality, which are particularly present after stroke. Therefore, to extend the applicability of motor unit-based clinical analyses, future work must continue to improve HD-EMG sensors, extend investigations to intramuscular EMG, and enhance decomposition techniques to increase motor unit yields across wider populations.

The synchronous behavior of motor units may be facilitated by preserved neural pathways in the higher functioning individuals. Recent studies have investigated altered behavior of motor unit and how the altered behavior correlated with functional impairments (e.g., Fugl-Meyer, fatigue, walking speed) (Kallenberg & Hermens, 2009; Hu *et al*., 2016; McManus *et al*., 2017; Negro *et al*., 2020; Li *et al*., 2024; Tacca *et al*., 2026). However, only Li et al. (2024) evaluated longitudinal relationships between motor unit number estimates/ F-waves and upper extremity Fugl-Meyer score, highlighting a critical next step in elucidating the neural mechanisms underlying the impaired motor control observed in this study. Future work should also combine recording modalities to evaluate changes in motor function at multiple levels (e.g., peripheral, spinal, brainstem, cortical) to more effectively map the altered neural networks responsible for post-stroke impaired balance.

## Data availability

Data can be made available upon reasonable request.

## Additional Information

## Author Contributions

Experiments were performed at Shirley Ryan AbilityLab. Levine, Jones, Avrillon, and Pons contributed to conception and design of the work. Levine, Yu, Jones, Muñoz, Avrillon, and Pons contributed to acquisition of data. Levine, Yu, Zaback, Thompson, Farina, Avrillon, and Pons contributed to analysis and interpretation of the data. All authors contributed to drafting and revising the manuscript. Additionally, all authors approved the final version of the manuscript; agree to be accountable for all aspects of the work in ensuring that questions related to the accuracy or integrity of any part of the work are appropriately investigated and resolved; and qualify for authorship and all those who qualify for authorship are listed.

## Acknowledgments

n/a

## References

Aman JE, Elangovan N, Yeh IL & Konczak J. (2014). The effectiveness of proprioceptive training for improving motor function: a systematic review. Front Hum Neurosci 8, 1075.

Arya KN & Pandian S. (2014). Interlimb neural coupling: Implications for poststroke hemiparesis. Annals of physical and rehabilitation medicine 57, 696–713.

Avrillon S, Hug F, Baker SN, Gibbs C & Farina D. (2024). Tutorial on MUedit: An open-source software for identifying and analysing the discharge timing of motor units from electromyographic signals. Journal of Electromyography and Kinesiology 77, 102886.

Beauchamp JA, Hassan AS, Mcpherson LM, Negro F, Pearcey GEP, Cummings M, Heckman C & Dewald JPA. (2023). Motor unit firing rate modulation is more impaired during flexion synergy-driven contractions of the biceps brachii in chronic stroke. Cold Spring Harbor Laboratory.

Berens P. (2009). : A MATLAB Toolbox for Circular Statistics. Journal of statistical software 31.

Boonstra TW, Daffertshofer A, Roerdink M, Flipse I, Groenewoud K & Beek PJ. (2009). Bilateral motor unit synchronization of leg muscles during a simple dynamic balance task. European Journal of Neuroscience 29, 613–622.

Brott T, Adams HP, Olinger CP, Marler JR, Barsan WG, Biller J, Spilker J, Holleran R, Eberle R & Hertzberg V. (1989). Measurements of acute cerebral infarction: a clinical examination scale. Stroke 20, 864–870.

Campbell BCV & Khatri P. (2020). Stroke. The lancet 396, 129–142.

Castronovo AM, Negro F, Conforto S & Farina D. (2015). The proportion of common synaptic input to motor neurons increases with an increase in net excitatory input. J Appl Physiol (1985) 119, 1337–1346.

Chia FS, Kuys S & Low Choy N. (2019). Sensory retraining of the leg after stroke: systematic review and meta-analysis. Clin Rehabil 33, 964–979.

Chou L-W, Palmer JA, Binder-Macleod S & Knight CA. (2013). Motor unit rate coding is severely impaired during forceful and fast muscular contractions in individuals post stroke. Journal of neurophysiology / 109, 2947–2954.

Cohen JW, Vieira T, Ivanova TD, Cerone GL & Garland SJ. (2021). Maintenance of standing posture during multi-directional leaning demands the recruitment of task-specific motor units in the ankle plantarflexors. Experimental Brain Research 239, 2569–2581.

Cohen JW, Vieira TM, Ivanova TD & Garland SJ. (2023a). Differential behavior of distinct motoneuron pools that innervate the triceps surae. Journal of neurophysiology / 129, 272–284.

Cohen JW, Vieira TM, Ivanova TD & Garland SJ. (2023b). Regional recruitment and differential behavior of motor units during postural control in older adults. Journal of neurophysiology / 130, 1321–1333.

Day BL, Steiger MJ, Thompson PD & Marsden CD. (1993). Effect of vision and stance width on human body motion when standing: implications for afferent control of lateral sway. J Physiol 469, 479–499.

Del Vecchio A, Holobar A, Falla D, Felici F, Enoka RM & Farina D. (2020). Tutorial: Analysis of motor unit discharge characteristics from high-density surface EMG signals. Journal of Electromyography and Kinesiology 53, 102426.

Di Giulio I, Maganaris CN, Baltzopoulos V & Loram ID. (2009). The proprioceptive and agonist roles of gastrocnemius, soleus and tibialis anterior muscles in maintaining human upright posture. The journal of physiology 587, 2399–2416.

Dias CP, Freire B, Goulart NBA, Dias De Castro C, Lemos FA, Becker J, Arndt A & Vaz MA. (2019). Impaired mechanical properties of Achilles tendon in spastic stroke survivors: an observational study. Top Stroke Rehabil 26, 261–266.

Dickstein R & Abulaffio N. (2000). Postural sway of the affected and nonaffected pelvis and leg in stance of hemiparetic patients. Archives of Physical Medicine and Rehabilitation 81, 364–367.

Dietz V & Schrafl-Altermatt M. (2016). Control of functional movements in healthy and post-stroke subjects: Role of neural interlimb coupling. Clinical neurophysiology : official journal of the International Federation of Clinical Neurophysiology 127, 2286–2293.

Garland S, Willems DA, Ivanova TD & Miller KJ. (2003). Recovery of standing balance and functional mobility after stroke11No commercial party having a direct financial interest in the results of the research supporting this article has or will confer a benefit upon the author(s) or upon any organization with which the author(s) is/are associated. Archives of physical medicine and rehabilitation 84, 1753–1759.

Garland SJ, Ivanova TD & Mochizuki G. (2007). Recovery of Standing Balance and Health-Related Quality of Life After Mild or Moderately Severe Stroke. Archives of physical medicine and rehabilitation 88, 218–227.

Garland SJ, Pollock CL & Ivanova TD. (2014). Could motor unit control strategies be partially preserved after stroke? Front Hum Neurosci 8, 864.

Gibbs J, Harrison LM & Stephens JA. (1995). Organization of inputs to motoneurone pools in man. The journal of physiology 485, 245–256.

Gorst T, Rogers A, Morrison SC, Cramp M, Paton J, Freeman J & Marsden J. (2019). The prevalence, distribution, and functional importance of lower limb somatosensory impairments in chronic stroke survivors: a cross sectional observational study. Disabil Rehabil 41, 2443–2450.

Granacher U, Bridenbaugh SA, Muehlbauer T, Wehrle A & Kressig RW. (2011). Age-related effects on postural control under multi-task conditions. Gerontology 57, 247–255.

Holobar A, Minetto MA & Farina D. (2014). Accurate identification of motor unit discharge patterns from high-density surface EMG and validation with a novel signal-based performance metric. Journal of neural engineering 11, 016008.

Hu X, Suresh AK, Rymer WZ & Suresh NL. (2015). Assessing altered motor unit recruitment patterns in paretic muscles of stroke survivors using surface electromyography. J Neural Eng 12, 066001.

Hu X, Suresh AK, Rymer WZ & Suresh NL. (2016). Altered motor unit discharge patterns in paretic muscles of stroke survivors assessed using surface electromyography. J Neural Eng 13, 046025.

Hug F, Avrillon S, Del Vecchio A, Casolo A, Ibanez J, Nuccio S, Rossato J, Holobar A & Farina D. (2021a). Analysis of motor unit spike trains estimated from high-density surface electromyography is highly reliable across operators. Journal of Electromyography and Kinesiology 58, 102548.

Hug F, Del Vecchio A, Avrillon S, Farina D & Tucker K. (2021b). Muscles from the same muscle group do not necessarily share common drive: evidence from the human triceps surae. Journal of applied physiology 130, 342–354.

Héroux ME, Dakin CJ, Luu BL, Inglis JT & Blouin J-S. (2014). Absence of lateral gastrocnemius activity and differential motor unit behavior in soleus and medial gastrocnemius during standing balance. Journal of applied physiology 116, 140–148.

Johnson CA, Biswas P, Tapia R, See J, Dodakian L, Chan V, Wang PT, Nenadic Z, Do AH & Reinkensmeyer DJ. (2025). The Weak Relationship Between Ankle Proprioception and Gait Speed After Stroke: A Robotic Assessment Study. Neurorehabil Neural Repair, 15459683251369497.

Kallenberg LA & Hermens HJ. (2009). Motor unit properties of biceps brachii in chronic stroke patients assessed with high-density surface EMG. Muscle Nerve 39, 177–185.

Kallenberg LAC & Hermens HJ. (2011). Motor unit properties of biceps brachii during dynamic contractions in chronic stroke patients. Muscle & Nerve 43, 112–119.

Karbasforoushan H, Cohen-Adad J & Dewald JPA. (2019). Brainstem and spinal cord MRI identifies altered sensorimotor pathways post-stroke. Nature Communications 10.

Katoozian L, Tahan N, Zoghi M & Bakhshayesh B. (2018). The Onset and Frequency of Spasticity After First Ever Stroke. J Natl Med Assoc 110, 547–552.

Khan F & Chevidikunnan MF. (2021). Prevalence of Balance Impairment and Factors Associated with Balance among Patients with Stroke. A Cross Sectional Retrospective Case Control Study. Healthcare 9, 320.

Kim H-Y. (2013). Statistical notes for clinical researchers: assessing normal distribution (2) using skewness and kurtosis. Restorative dentistry & endodontics 38, 52.

Li X, Holobar A, Gazzoni M, Merletti R, Rymer WZ & Zhou P. (2015). Examination of Poststroke Alteration in Motor Unit Firing Behavior Using High-Density Surface EMG Decomposition. IEEE Trans Biomed Eng 62, 1242–1252.

Li X, Shao Z, Li Z, Wei X, Zong L, Wang P, Zhou T & Wang H. (2024). The relationship between the functional status of upper extremity motor neurons and motor function and prognosis in stroke patients. Frontiers in Neurology 15.

Li X, Wang Y-C, Suresh NL, Rymer WZ & Zhou P. (2011). Motor Unit Number Reductions in Paretic Muscles of Stroke Survivors. IEEE Transactions on Information Technology in Biomedicine 15, 505–512.

Liang JN, Bashford G, Kulig K & Ho KY. (2024). Achilles tendon morphology adaptations in chronic post-stroke hemiparesis: a comparative analysis with neurologically intact controls. Front Sports Act Living 6, 1498333.

Loram ID, Maganaris CN & Lakie M. (2004). Paradoxical muscle movement in human standing. The journal of physiology 556, 683–689.

Loram ID, Maganaris CN & Lakie M. (2005). Human postural sway results from frequent, ballistic bias impulses by soleus and gastrocnemius. The journal of physiology 564, 295–311.

Lukács M. (2005). Electrophysiological signs of changes in motor units after ischaemic stroke. Clinical neurophysiology : official journal of the International Federation of Clinical Neurophysiology 116, 1566–1570.

Lundström E, Smits A, Terént A & Borg J. (2010). Time-course and determinants of spasticity during the first six months following first-ever stroke. J Rehabil Med 42, 296–301.

Marsden JF. (2005). The vestibular control of balance after stroke. *Journal of Neurology*, Neurosurgery & Psychiatry 76, 670–679.

Matsuda K, Murai I, Okoba R, Ikeda T & Takano Y. (2025). Differences in Postural Control Associated With Aging and Executive Function. Cureus 17, e81771.

Mccall AA, Miller DM & Yates BJ. (2017). Descending Influences on Vestibulospinal and Vestibulosympathetic Reflexes. Frontiers in Neurology 8.

McManus L, Hu X, Rymer WZ, Suresh NL & Lowery MM. (2017). Motor Unit Activity during Fatiguing Isometric Muscle Contraction in Hemispheric Stroke Survivors. Front Hum Neurosci 11, 569.

McNulty PA, Lin G & Doust CG. (2014). Single motor unit firing rate after stroke is higher on the less-affected side during stable low-level voluntary contractions. Front Hum Neurosci 8, 518.

Miller DM, Klein CS, Suresh NL & Rymer WZ. (2014a). Asymmetries in vestibular evoked myogenic potentials in chronic stroke survivors with spastic hypertonia: Evidence for a vestibulospinal role. Clinical Neurophysiology 125, 2070–2078.

Miller LC, Thompson CK, Negro F, Heckman CJ, Farina D & Dewald JP. (2014b). High-density surface EMG decomposition allows for recording of motor unit discharge from proximal and distal flexion synergy muscles simultaneously in individuals with stroke. Annu Int Conf IEEE Eng Med Biol Soc 2014, 5340–5344.

Mizrahi J, Solzi P, Ring H & Nisell R. (1989). Postural stability in stroke patients: Vectorial expression of asymmetry, sway activity and relative sequence of reactive forces. Medical & Biological Engineering & Computing 27, 181–190.

Mochizuki G, Ivanova TD & Garland SJ. (2007). Factors affecting the common modulation of bilateral motor unit discharge in human soleus muscles. J Neurophysiol 97, 3917–3925.

Monsour M, Ivanova TD, Wilson TD & Garland SJ. (2012). Influence of Vestibular Afferent Input on Common Modulation of Human Soleus Motor Units During Standing. Motor Control 16, 466–479.

Mottram CJ, Heckman CJ, Powers RK, Rymer WZ & Suresh NL. (2014). Disturbances of motor unit rate modulation are prevalent in muscles of spastic-paretic stroke survivors. J Neurophysiol 111, 2017–2028.

Murphy SA, Negro F, Farina D, Onushko T, Durand M, Hunter SK, Schmit BD & Hyngstrom A. (2018). Stroke increases ischemia-related decreases in motor unit discharge rates. J Neurophysiol 120, 3246–3256.

Negro F, Bathon KE, Nguyen JN, Bannon CG, Orizio C, Hunter SK & Hyngstrom AS. (2020). Impaired Firing Behavior of Individually Tracked Paretic Motor Units During Fatiguing Contractions of the Dorsiflexors and Functional Implications Post Stroke. Frontiers in neurology 11, 540893.

Negro F, Muceli S, Castronovo AM, Holobar A & Farina D. (2016). Multi-channel intramuscular and surface EMG decomposition by convolutive blind source separation. J Neural Eng 13, 026027.

Palmer JA, Payne AM, Mirdamadi JL, Ting LH & Borich MR. (2025). Delayed Cortical Responses During Reactive Balance After Stroke Associated With Slower Kinetics and Clinical Balance Dysfunction. Neurorehabilitation & neural repair 39, 16–30.

Pollock CL, Ivanova TD, Hunt MA & Garland SJ. (2014). Motor unit recruitment and firing rate in medial gastrocnemius muscles during external perturbations in standing in humans. Journal of neurophysiology / 112, 1678–1684.

Promsri A. (2022). Modulation of bilateral lower-limb muscle coordination when performing increasingly challenging balance exercises. Neuroscience letters 767, 136299.

Tacca N, Levine JT, Heimann MK, Schlink BR, Cady S, Colachis SC, Baumgart I, Bollinger A, Dunlap C, Putnam P, Darrow MJ, Wengerd L, Pons JL, Friedenberg DA & Meyers EC. (2026). Leveraging neural drive to assess hand motor function in individuals with chronic stroke. J Neuroeng Rehabil 23, 7.

Tamura S, Miyata K, Kobayashi S, Takeda R & Iwamoto H. (2022). The minimal clinically important difference in Berg Balance Scale scores among patients with early subacute stroke: a multicenter, retrospective, observational study. Top Stroke Rehabil 29, 423–429.

Thomas CK, Butler JE & Zijdewind I. (2002). Patterns of pathological firing in human motor units. Adv Exp Med Biol 508, 237–244.

Thompson CK, Negro F, Johnson MD, Holmes MR, Mcpherson LM, Powers RK, Farina D & Heckman CJ. (2018). Robust and accurate decoding of motoneuron behaviour and prediction of the resulting force output. The Journal of Physiology 596, 2643–2659.

Ting LH. (2007). Dimensional reduction in sensorimotor systems: a framework for understanding muscle coordination of posture. In Progress in Brain Research, pp. 299–321. Elsevier.

Vieira TMM, Loram ID, Muceli S, Merletti R & Farina D. (2012). Recruitment of motor units in the medial gastrocnemius muscle during human quiet standing: is recruitment intermittent? What triggers recruitment? Journal of neurophysiology / 107, 666–676.

Welch TDJ & Ting LH. (2014). Mechanisms of Motor Adaptation in Reactive Balance Control. PLoS ONE 9, e96440.

West SG, Finch JF & Curran PJ. (1995). Structural equation models with nonnormal variables: Problems and remedies. In Structural Equation Modeling: Concepts, Issues, and Applications, ed. Hyole RH, pp. 56–75. Sage Publication, Inc Thousand Oaks, CA, US.

Winter D, Prince F, Frank J, Powell C & Zabjek K. (1996). Unified theory regarding A/P and M/L balance in quiet stance Journal of Neurophysiology 75, 2334–2343.

Winter DA, Patla AE, Ishac M & Gage WH. (2003). Motor mechanisms of balance during quiet standing. Journal of electromyography and kinesiology : official journal of the International Society of Electrophysiological Kinesiology 13, 49–56.

Winter DA, Prince F, Stergiou P & Powell C. (1993). Medial-lateral and anterior-posterior motor responses associated with centre of pressure changes in quiet standing. Neuroscience Research Communications 12, 141–148.

Wutzke CJ, Mercer VS & Lewek MD. (2013). Influence of lower extremity sensory function on locomotor adaptation following stroke: a review. Top Stroke Rehabil 20, 233–240.

Yamanaka E, Goto R, Kawakami M, Tateishi T, Kondo K & Nojima I. (2023). Intermuscular Coherence during Quiet Standing in Sub-Acute Patients after Stroke: An Exploratory Study. Brain Sciences 13, 1640.

Zaback M, Missen KJ, Adkin AL, Chua R, Inglis JT & Carpenter MG. (2023). Cortical potentials time-locked to discrete postural events during quiet standing are facilitated during postural threat exposure. The Journal of Physiology 601, 2473–2492.

Zaback M & Thompson CK. (2026). Independent directional tuning of the human triceps surae muscles during standing postural control. J Appl Physiol (1985).

Zhao H, Ren Y, Roth EJ, Harvey RL & Zhang LQ. (2015). Concurrent deficits of soleus and gastrocnemius muscle fascicles and Achilles tendon post stroke. J Appl Physiol (1985) 118, 863–871.

Zhao H, Ren Y, Wu YN, Liu SQ & Zhang LQ. (2009). Ultrasonic evaluations of Achilles tendon mechanical properties poststroke. J Appl Physiol (1985) 106, 843–849.

